# Genetic risk and immune dysregulation of classic Hodgkin lymphoma transformation of chronic lymphocytic leukemia/small lymphocytic lymphoma: a multicentric study

**DOI:** 10.64898/2026.05.11.26352584

**Authors:** Mingfei Yan, Sameer A. Parikh, Michelly K. Sampaio De Melo, Paul J. Hampel, Nathaniel Aleynick, Alexander Chan, Ozgur Can Eren, Katherine Lopez, Alexa Cohen, Mikhail Roshal, Megan S. Lim, Leonardo Boiocchi, Ahmet Dogan, Yanming Zhang, Sutapa Sinha, Kari G. Rabe, Neil E. Kay, Elaine S. Jaffe, Rebecca L. King, Wenbin Xiao

**Author notes:** **Corresponding author:** Wenbin Xiao, MD, PhD. These authors contribute equally (Sameer A. Parikh, Michelly K. Sampaio De Melo). These authors contribute equally (Elaine S. Jaffe, Rebecca L. King). Department of Pathology and Laboratory Medicine, H. Lee Moffitt Cancer Center & Research Institute, Tampa, FL. **Author contributions:** M.Y., S.A.P., M.S.D.M., E.S.J., R.K., and W.X. collected and reviewed cases. P.J.H., K.L., A. Cohen, S.S., K.G.R., and N.E.K. collected data. N.A., M.S.L., L.B., and A.D. performed and analyzed multiplexed immunofluorescence data. A. Chan., O.C.E., and M.R. performed and analyzed flow cytometric data. Y.Z. performed and analyzed FISH studies. M.Y. and W.X. analyzed all the data and drafted the manuscript. W.X. conceived and supervised the study and provided research support. **Data-sharing statement:** Original data can be shared upon request by contacting the corresponding author.

## Abstract

Richter transformation of Chronic lymphocytic leukemia/small lymphocytic lymphoma (CLL/SLL) into classic Hodgkin lymphoma (CHL-RT) is rare and remains incompletely understood. Two histologic subtypes are recognized: type 1 (CLL/SLL with scattered Hodgkin/Reed-Sternberg (HRS) cells) and type 2 (HRS cells within a polymorphous inflammatory background). In this multi-institutional study of 77 patients with CHL-RT (27 type 1 and 50 type 2), we characterized immune evasion markers, *PD-L1*/*PD-L2* copy number alterations, tumor microenvironment, and performed targeted next-generation sequencing on 37 CLL/SLL samples. HRS cells in CHL-RT displayed immune-evasion phenotypes similar to de novo CHL, though PD-L1 expression was lower in type 1 cases. *PD-L1/PD-L2* gain/polysomy were frequent (83.3%). CLL/SLL with CHL-RT harbored increased mutations in *XPO1, FBXW7, BIRC3, TRAF3*, and *HLA-A* versus reference CLL/SLL. Similar mutational profiles, demographics, and survival outcomes support a biological continuum between type 1 and type 2 CHL-RT, with distinct genetic features in CLL/SLL predisposing to CHL transformation.

**STATEMENT OF SIGNIFICANCE:** Classic Hodgkin lymphoma-type Richter transformation arises from convergent Hodgkin-like immune evasion on a genetically primed CLL/SLL background. Through integrated genomic and tumor microenvironment analyses in a large multicenter cohort, we establish this as a biologically coherent entity and provide rationale for PD-L1/PD-L2-directed therapeutic strategies.

## INTRODUCTION

Large cell transformation of chronic lymphocytic leukemia/small lymphocytic lymphoma (CLL/SLL), known as Richter’s transformation (RT), frequently manifests as diffuse large B-cell lymphoma (DLBCL-RT) and occasionally exhibits features of classic Hodgkin lymphoma (CHL-RT). The incidence of CHL-RT is approximately 0.4% to 0.7% among CLL/SLL patients(1,2). CHL-RT typically develops several years after the initial diagnosis of CLL/SLL, with a median interval ranging from 4.3 to 5.9 years(3,4). Clinically, patients with CHL-RT generally experience poor outcomes, with reported median overall survival ranging from 1.1 to 3.9 years following the diagnosis of CHL (2,5-7). However, patients treated with CHL-directed regimens have demonstrated more favorable outcomes, with median overall survival extending from 4.8 to 13.2 years (6,8).

CHL-RT is characterized by the presence of Hodgkin/Reed-Sternberg (HRS) cells that share morphologic and immunophenotypic features with those seen in *de novo* CHL. CHL-RT can present in two distinct histologic patterns: type 1 (CLL with HRS cells), which features scattered HRS cells in a background of CLL/SLL but lacks the polymorphous inflammatory microenvironment typically seen in CHL; and type 2 (CLL with CHL), which demonstrates a full manifestation of CHL, including the characteristic inflammatory background. Previous studies have suggested that HRS cells in both type 1 and type 2 patients exhibit similar immunophenotypes and frequently demonstrate EBV-encoded RNA (EBER) positivity in 60-70% of cases(6,9). Furthermore, HRS cells in both types of CHL-RT can be clonally related to the underlying CLL/SLL(9,10). Importantly, type 1 may progress to type 2, and both types exhibit similar outcomes(9). In addition, type 1 patients show improved outcomes when treated with CHL-directed compared to CLL-directed therapies(6). These findings suggest that both type 1 and type 2 CHL-RT likely represent a spectrum of the same process with type 1 representing an earlier stage.

While HRS cells in CHL-RT exhibit a characteristic CHL immunophenotype, as demonstrated by diagnostic markers such as CD20, PAX5, CD30, and CD15, it remains unclear whether these cells also display dysregulation of markers associated with immune evasion, a hallmark of *de novo* CHL. These include gain or amplification of *PD-L1/PD-L2* genes, overexpression of PD-L1/PD-L2 proteins, and downregulation of molecules involved in antigen presentation(11-13). In addition, HRS cells in *de novo* CHL demonstrate a distinct tumor microenvironment (TME), characterized by enrichment of cytotoxic T-lymphocyte-associated protein 4 (CTLA-4)-positive inhibitory T cells (14). Understanding these aspects may provide critical insights into the pathogenesis of CHL-RT.

Genetic factors may predispose patients with CLL/SLL to RT. DLBCL-RT is genetically characterized by mutations in several key genes, including *TP53* disruption(15-17), *CDKN2A/B* inactivation(16,18), *NOTCH1* mutations(16,17), and *MYC* activation(15-17), which collectively suggest a higher risk of DLBCL-RT development in CLL/SLL patients harboring these alterations. *De novo* CHL, on the other hand, is defined by mutations and copy number alterations affecting various biological pathways, such as activation of JAK-STAT and NF-κB signaling, immune evasion, and disruption of cellular homeostasis through *XPO1* mutation(19,20). In contrast, the genetic features of CLL/SLL associated with CHL-RT have not been studied.

Leveraging a large cohort of patients with CHL-RT from 3 academic institutions, this study aims to deepen our understanding of its underlying biology. We identified a shared immune evasion phenotype in HRS cells between both types of CHL-RT and *de novo* CHL. We observed a similar TME enriched for CTLA-4-positive T cells in type 2 CHL-RT and *de novo* CHL, which was less pronounced in HRS cells from type 1 CHL-RT. Finally, we discovered distinct mutational profiles in CLL/SLL with CHL-RT, including frequent mutations in *XPO1* and *FBXW7*, immunoglobulin gene (*HLA-A*), and genes in the NF-κB pathway (*BIRC3* and *TRAF3*).

## RESULTS

### HRS cells in CHL-RT show immune evasion immunophenotype and frequent EBV positivity

We compared the immunophenotype of HRS cells between type 1 and type 2 CHL-RT, and against de novo CHL (**Figures 1B, 2A, and 2B**). HRS cells from both groups exhibited a diagnostic CHL immunophenotype, including uniform CD30 positivity, frequent CD15 expression (57.7% vs. 72%, p=0.30), dim PAX5 expression (87% vs. 81.4%, p=1.00), and frequent negativity for CD20 (72.2% vs. 78.6%, p=0.74) and CD45 (100% vs. 90.9%, p=0.55). EBV positivity, as detected by EBER staining, was observed in 62.5% of type 1 and 63.2% of type 2 CHL-RT cases (p=1), both significantly higher than in de novo CHL (17.2%, p=0.001 and p<0.001, respectively) (**Figure 1C**). Beyond diagnostic markers, HRS cells in both CHL-RT types frequently demonstrated an immune-evasive immunophenotype, evidenced by PD-L1 expression, loss of antigen-presentation molecules, and MHC-II negativity. Notably, PD-L1 expression was significantly lower in type 1 CHL-RT (53.3%) compared to both type 2 CHL-RT (100%, p<0.001) and de novo CHL (85.7%, p=0.009). MHC-II loss was more frequent in type 1 (42.9%) and type 2 (48.2%) CHL-RT than in de novo CHL (21.3%, p=0.16 and p=0.02, respectively), while MHC-I (18.2% vs. 30%, p=0.68), B2M (40% vs. 40%, p=1.0), and CIITA (7.1% vs. 7.1%, p=1.0) loss was comparable across both CHL-RT types. PD-L2 expression was absent in type 1 and detected in only 10% of type 2 CHL-RT cases (p=0.51), with no significant differences observed compared to de novo CHL.

**Figure 1.**
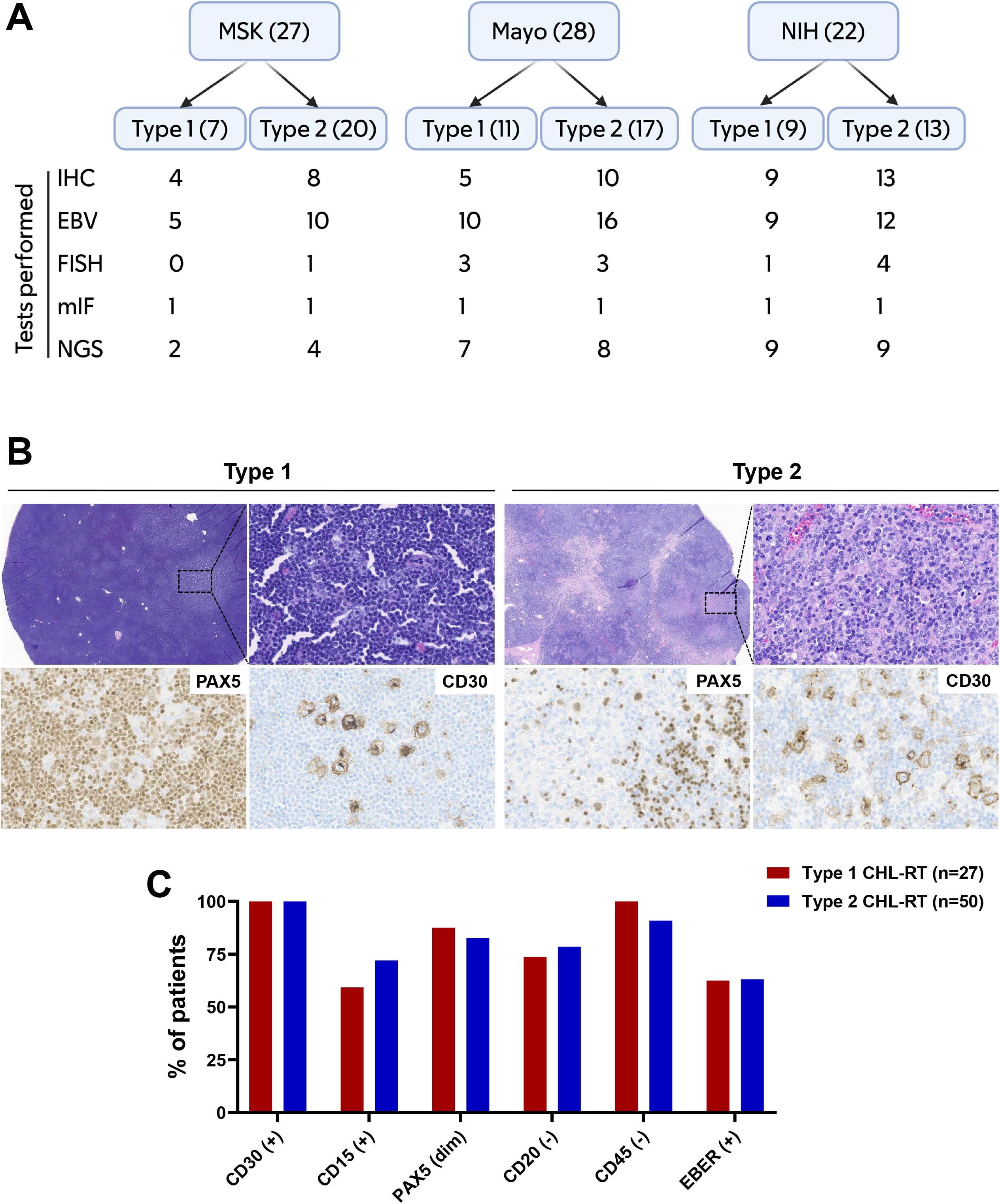
Cohort composition and baseline immunophenotype of Hodgkin/Reed-Sternberg (HRS) cells in classical Hodgkin lymphoma-type Richter transformation (CHL-RT). (A) Summary of case contributions from three institutions and the number of cases included in downstream immunophenotypic, tumor microenvironment, and molecular genetic analyses (MSK, Memorial Sloan Kettering Cancer Center; Mayo, Mayo Clinic Rochester; NIH, National Institutes of Health/National Cancer Institute). (B) Representative histopathologic features of type 1 and type 2 CHL□ RT. Type 1 CHL□ RT demonstrates scattered HRS cells (highlighted by CD30 IHC) within a chronic lymphocytic leukemia/small lymphocytic lymphoma (CLL/SLL) background (highlighted by PAX5 IHC), whereas type 2 CHL□ RT shows HRS cells embedded in a characteristic polymorphous inflammatory background. (C) Comparison of baseline immunophenotypic features and Epstein-Barr virus (EBV) status of HRS cells, assessed by EBER in situ hybridization, between type 1 and type 2 CHL□ RT. ns, not statistically significant.

**Figure 2.**
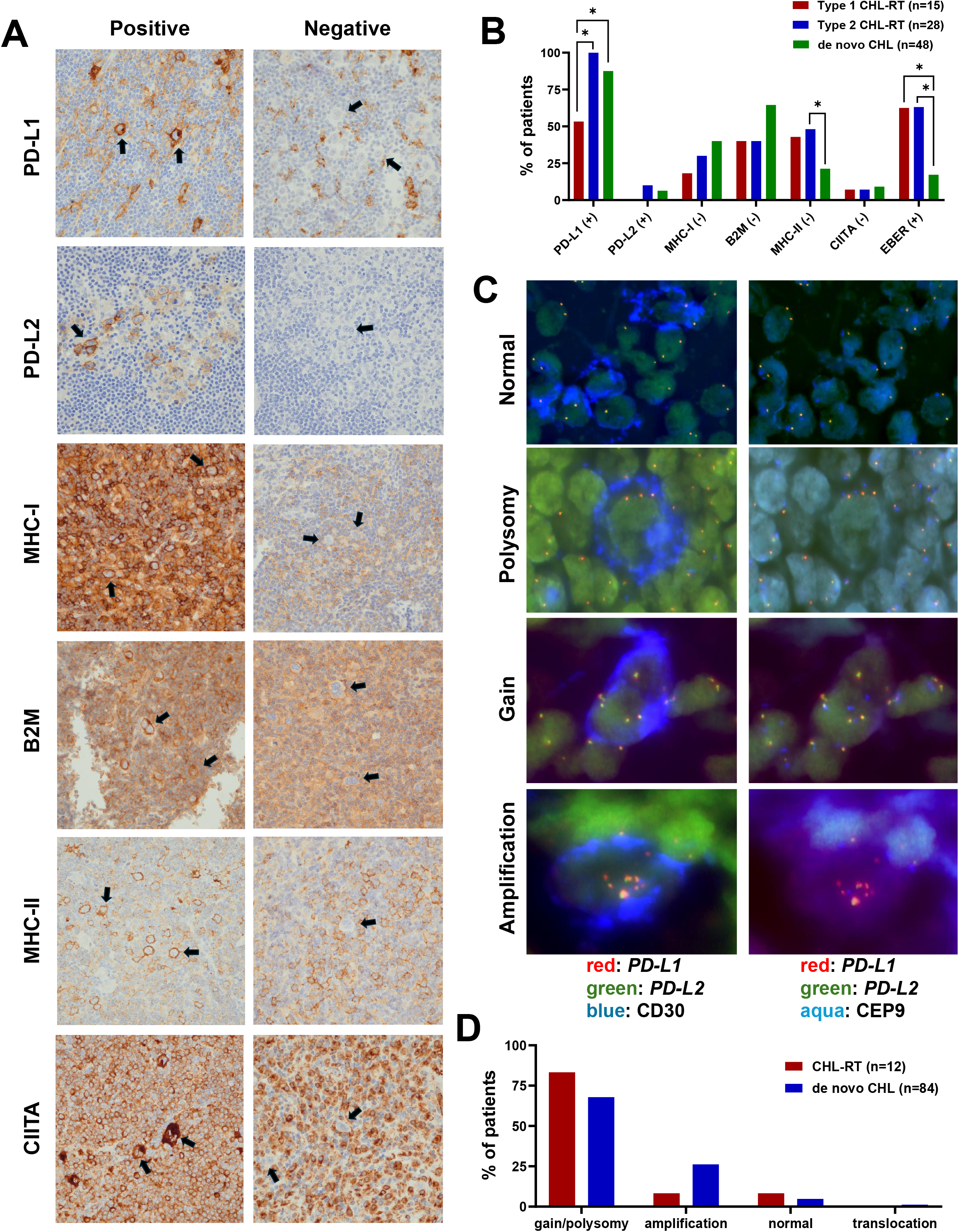
Immune evasion phenotype and PD□L1/PD□L2 copy number alterations in CHL□RT compared with *de novo* CHL. (A) Representative immunohistochemical images illustrating immune evasion marker expression patterns in CHL-RT HRS cells (positive examples shown on the left, negative examples on the right), including PD□ L1, PD□ L2, MHC□ I, B2M, MHC□ II, and CIITA. Representative HRS cells are indicated by black arrows. (B) Comparison of immune evasion marker expression, along with EBV status by EBER in situ hybridization, in HRS cells from type 1 CHL-RT, type 2 CHL-RT, and *de novo* CHL. *, p < 0.05. (C) Assessment of *PD*□*L1/PD*□*L2* copy number alterations (CNAs) in CHL-RT using combined CD30 immunofluorescence (IF) and *PD□L1/PD□L2* fluorescence in situ hybridization (FISH) with CEP9 (centromeric probe for chromosome 9) as a control. Representative images demonstrate patterns classified as normal, polysomy, gain, and amplification of *PD*□*L1/PD*□*L2* (definitions provided in Materials and Methods). For each field, paired images are shown, with the left panel displaying *PD*□*L1, PD*□*L2*, and CD30 signals, and the right panel displaying *PD*□*L1, PD*□*L2*, and CEP9 signals. (D) Comparison of *PD□L1/PD*□*L2* CNAs in HRS cells from CHL□ RT versus *de novo* CHL.

### HRS cells in CHL-RT show frequent gain or polysomy of *PD-L1/PD-L2*

We examined *PD-L1/PD-L2* copy number alterations in HRS cells from 12 CHL-RT cases (four cases of type 1 and eight cases of type 2) (**Figure 2C**) and compared these findings to a cohort of 84 *de novo* CHL cases diagnosed at our institution since 2021. Compared to *de novo* CHL, HRS cells in CHL-RT numerically exhibited a higher frequency of *PD-L1/PD-L2* gain or polysomy (83.3% vs. 67.9%, p=0.34), a lower frequency of *PD-L1/PD-L2* amplification (8.3% vs. 26.2%, p=0.28), and similar low frequencies of cases with normal *PD-L1/PD-L2* copy numbers (8.3% vs. 4.8%, p=0.49). *PD-L1/PD-L2* translocation, observed in only one of the 84 *de novo* CHL cases, was not detected in HRS cells associated with CHL-RT (**Figure 2D**).

Apart from two type 1 cases with *PD-L1/PD-L2* polysomy that exhibited PD-L1 negativity by IHC, all other nine cases with available IHC results showed PD-L1 positivity. These included seven cases with *PD-L1/PD-L2* polysomy, one case with *PD-L1/PD-L2* amplification, and one case with normal *PD-L1/PD-L2* copy number.

### HRS cells have distinct immune microenvironment in type 1 vs 2 CHL-RT

mIF studies were performed to evaluate the TME of HRS cells in a subset of type 1 and type 2 CHL-RT, as well as in *de novo* CHL (**Figure 3A**). Using a limited panel of 6 markers, we primarily assessed the numbers of CTLA-4-positive and PD-1-positive T cells, as well as B cells, within a 75 µm radius of HRS cells (**Figure 3B**). HRS cells in both type 2 CHL-RT (median: 39 cells, p<0.001) and *de novo* CHL (median: 18 cells, p=0.002) demonstrated a similar enrichment of CTLA-4-positive T cells) when compared to the HRS cells in type 1 CHL-RT (median: 6 cells). The number of PD-1-positive T cells within the TME of HRS cells was generally lower than that of CTLA-4-positive T cells but was also more frequently observed in type 2 CHL-RT (median: 6 cells, p=0.041) and *de novo* CHL (median: 5 cells, p=0.092) than in type 1 CHL-RT (median: 2 cells). Finally, as expected, B cells (likely CLL cells) were significantly more abundant in the TME of HRS cells with type 1 CHL-RT (median: 101 cells) than those with type 2 CHL-RT (median: 34 cells, p=0.008) or in *de novo* CHL (median: 7 cells, p<0.001); the difference between type 2 CHL-RT and *de novo* CHL was also statistically significant (p=0.001).

**Figure 3.**
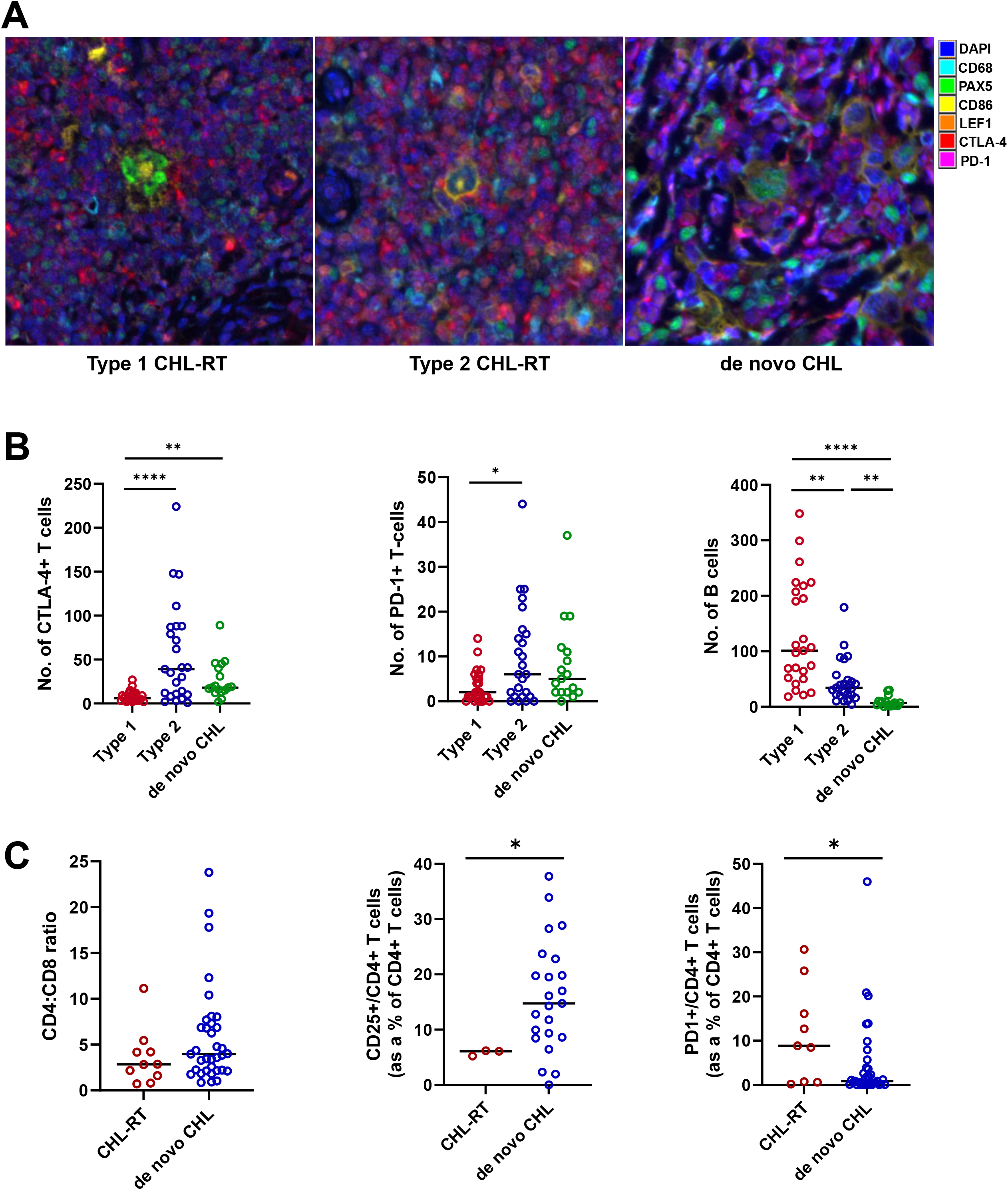
Spatial immune microenvironment of HRS cells in type 1 and type 2 CHL□RT and *de novo* CHL. (A) Representative multiplex immunofluorescence (mIF) images depicting the cellular neighborhood surrounding HRS cells in type 1 CHL-RT, type 2 CHL-RT, and *de novo* CHL. The six□marker panel includes CD68, PAX5, CD86, LEF1, CTLA-4, and PD-1, with DAPI nuclear counterstain. HRS cells are identified based on morphology and co□expression of PAX5 and CD86. (B) Quantitative spatial analysis of CTLA-4□ cells, PD□ 1□ cells, and B□cell populations within a 75□µm radius of individual HRS cells. A total of 25 HRS cells from three type 1 cases, 25 HRS cells from three type 2 cases, and 17 HRS cells from two *de novo* CHL cases were analyzed. Black bars indicate group medians. *, p < 0.05; **, p < 0.01; ****, p < 0.0001. (C) Flow cytometric comparison of T□ cell subsets between CHL□ RT and *de novo* CHL, including CD4:CD8 ratio, proportion of CD25□ T cells, and proportion of PD-1□ cells within the CD4□ T□ cell compartment. Black bars indicate group medians. *, p < 0.05.

We further compared T cell composition by flow cytometry between CHL-RT and a cohort of *de novo* CHL (**Figure 3C**). The two groups demonstrated similar CD4:CD8 ratios within the T cell compartment (median, 2.8 vs 4.0; *p*=0.196). In contrast, CHL-RT cases exhibited lower proportions of CD25□ T cells (median, 6.1% vs 14.8%; *p*=0.041) and higher proportions of PD-1□ T cells (median, 8.9% vs 0.9%; *p*=0.043) within the CD4□ T cell compartment compared with *de novo* CHL.

### CLL/SLL with CHL-RT show distinct genetic profiles

NGS studies were conducted on bulk FFPE tissue of 37 CLL/SLL samples with CHL-RT (HRS cells <5%), including 18 samples with type 1 and 19 samples with type 2 CHL-RT. Genes recurrently mutated in more than one sample were selected for further comparison **(Figure 4A**). The most frequently mutated genes in the combined cohort of type 1 and type 2 CHL-RT included *XPO1* (16.2%, 6/37), *FBXW7* (16.2%, 6/37), *BIRC3* (13.5%, 5/37), *SF3B1* (10.8%, 4/37), *NOTCH1* (10.8%, 4/37), and *TP53* (10.8%, 4/37), among others. Mutations in several genes were more frequently observed in type 1 CLL/SLL, including *XPO1* (27.8% vs 5.3%), *NOTCH1* (16.7% vs 5.3%), *BRAF* (16.7% vs 0%), *KRAS* (11.1% vs 0%), *HLA-A* (11.1% vs 0%), *EGR2* (11.1% vs 0%), *SETD2* (11.1% vs 0%), and *MED12* (11.1% vs 0%), while other genes were more frequently mutated in type 2 CLL/SLL, including *TP53* (5.6% vs 15.8%) and *CHD2* (0% vs 10.5%); however, these differences did not reach statistical significance (**Figure 4B**). Variant allele frequencies (VAFs) for all mutations were greater than 2%, with the majority exceeding 5% (**Figure 4C)**, suggesting that these alterations were unlikely to be derived from HRS cells.

**Figure 4.**
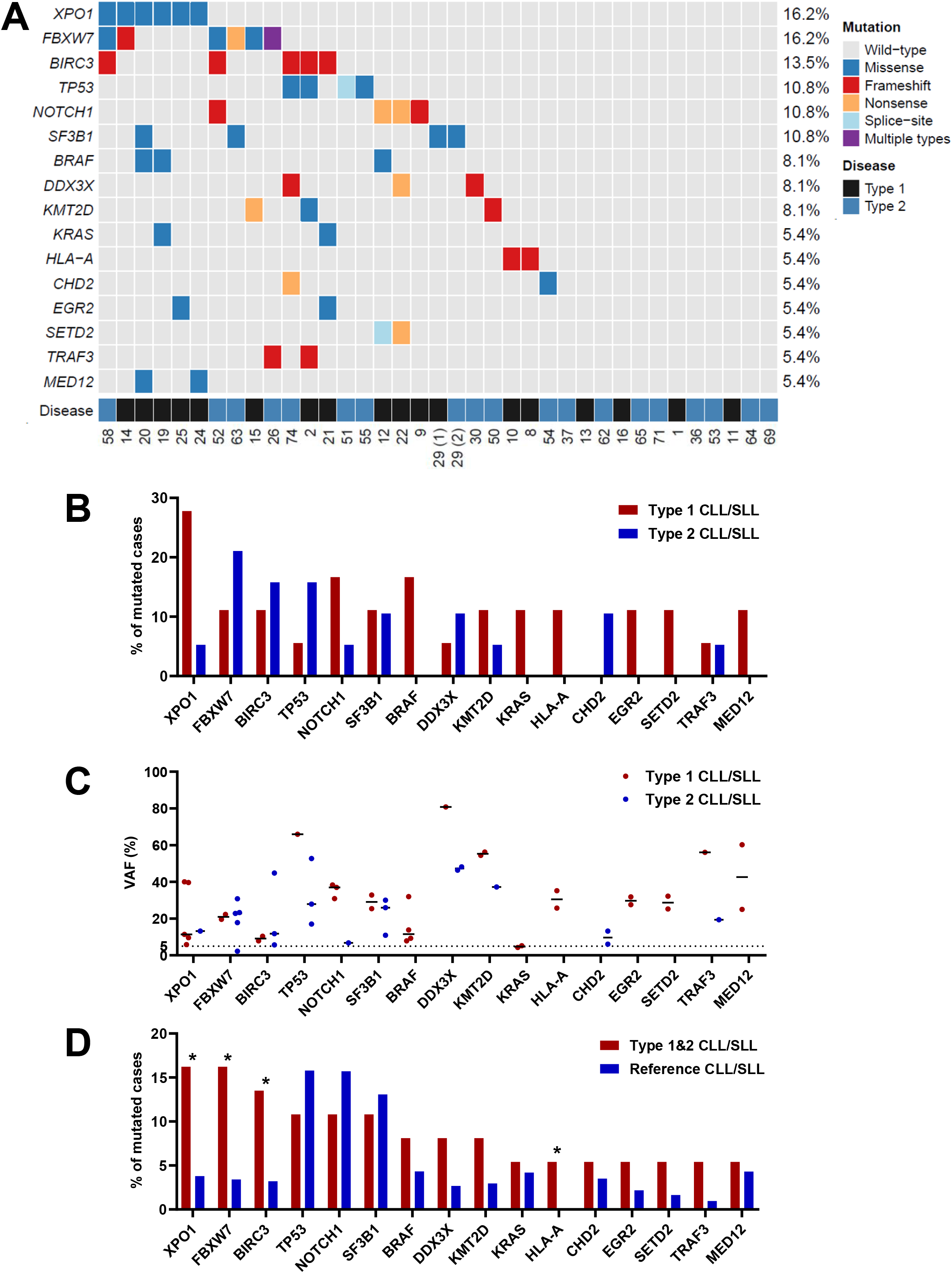
Distinct mutational landscape of CLL/SLL associated with CHL□RT. (A) Oncoplot summarizing pathogenic and likely pathogenic variants identified by targeted next□generation sequencing (MSK□ IMPACT□ Heme) in bulk CLL/SLL samples from patients with CHL-RT. Genes recurrently mutated in two or more samples are shown. CHL□ RT subtype (type 1 vs type 2) and de-identified patient IDs are indicated at the bottom of the plot. (B) Comparison of mutation frequencies between type 1 and type 2 CHL□ RT-associated CLL/SLL for selected recurrently mutated genes. Differences did not reach statistical significance and corresponding p values are not shown. (C) Comparison of variant allele frequencies between type 1 and type 2 CHL□ RT-associated CLL/SLL for selected recurrently mutated genes. Black bars indicate group median. (D) Comparison of mutation frequencies in CHL□ RT-associated CLL/SLL versus a reference CLL/SLL cohort derived from institutional databases and published cohorts. *, p < 0.05.

*XPO1* and *FBXW7*, two genes involved in maintaining protein homeostasis, were among the most frequent alterations (10 cases in total, 27%). All *XPO1* mutations were missense mutations at the hotspot locus p.E571. Mutations involving the epigenetic regulators (*KMT2D*/*CHD2*/*MED12*, 7 cases in total, 18.9%) or the canonical NF-κB pathway (*BIRC3*/*TRAF3*, 6 cases in total, 16.2%) were also frequently detected. Other involved pathways include Notch signaling (*NOTCH1/2*, 5 cases in total, 13.5%), RNA splicing (*SF3B1*, 4 cases, 10.8%), MAPK pathway signaling (*BRAF/KRAS*, 4 cases, 10.8%). Of note, three of the four patients with *TP53* mutations belonged to the type 2 CHL-RT group, and an *ATM* mutation was only detected in one type 2 CHL-RT case.

When comparing the combined CHL-RT cohort to a reference CLL/SLL cohort (n=2743), comprising cases from our in-house clinical sequencing database and data from three additional studies available through cBioPortal(21-23), several genes were found to be significantly more frequently mutated in CLL/SLL with CHL-RT (**Figure 4D**). These included *XPO1* (16.2% vs. 3.8%, p=0.002), *FBXW7* (16.2% vs. 3.4%, p=0.001), *BIRC3* (13.5% vs. 3.2%, p=0.006), and *HLA-A* (5.4% vs. 0.05%, p=0.001). *TRAF3* was also more commonly mutated in CLL/SLL with CHL-RT despite not reaching statistical significance (5.4% vs 1.0%, p=0.053).

One patient (#29) was initially diagnosed with type 1 CHL-RT and subsequently developed type 2 disease about six months later. NGS was performed on both CHL-RT samples and revealed identical mutation in *SF3B1* p.R775Q with similar VAFs (33% in type 1 vs 30% in type 2).

NGS was also performed on bulk FFPE tissue from three patients with type 2 CHL□ RT in whom the samples showed CHL with a high density of HRS cells and no morphologic or immunophenotypic evidence of the previously diagnosed CLL/SLL. The first patient (case #28), with approximately 10% HRS cells, harbored an *HLA*□*A* mutation (VAF 11.2%). The second patient (case #59), with 20–30% HRS cells, harbored an *SRSF2* mutation (VAF 10.7%). The third patient (case #68), with 10–20% HRS cells, had no pathogenic or likely pathogenic mutations detected.

### Comparison of mutational profiles in paired CHL and CLL/SLL

We conducted paired NGS on macrodissected areas enriched for either CLL/SLL or CHL from the same specimens in 3 type 2 CHL-RT patients, enabling us to analyze the mutational spectrum and clonal relationships between the two neoplastic processes (**Figure 5**). The proportion of CLL/SLL and HRS cells in the macrodissected areas was estimated by PAX5 (focusing on positive small cells) and CD30 IHCs, respectively. In the first patient, distinct mutations were identified in the CLL/SLL- and CHL-enriched areas, with no shared mutations observed. Given the neoplastic burden in each area, it is most likely that there were no shared mutations between the CLL/SLL and HRS cells in this case. The second patient exhibited multiple shared mutations with similar VAFs between the CLL/SLL- and CHL-enriched areas, indicating a clonal relatedness. Considering the relative abundance of HRS cells, it is favored that these mutations indeed originate from HRS cells, rather than background CLL/SLL. In the third patient, no mutation was detected in both the CLL/SLL and the CHL-enriched area; however, as HRS cells constituted only 3-5% of the macrodissected CHL area, the possibility of false negativity due to low tumor content cannot be excluded.

**Figure 5.**
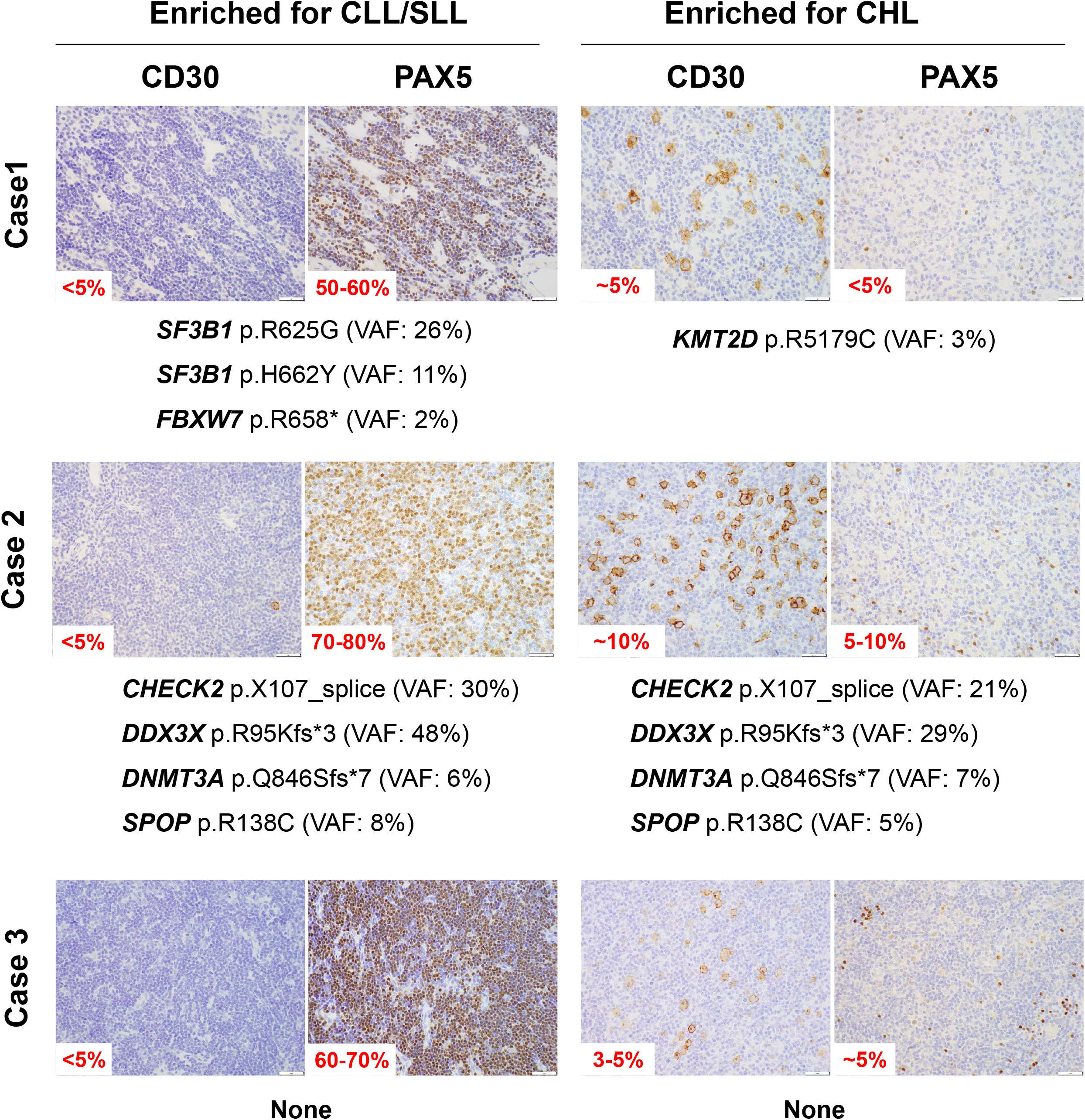
Paired sequencing of macrodissected CLL/SLL□ and CHL□enriched areas in type 2 CHL□RT. Targeted next□ generation sequencing (NGS) results from paired macrodissected regions enriched for CLL/SLL versus CHL from the same specimens in three patients with type 2 CHL□ RT. Estimated neoplastic burden in each region was assessed by PAX5 and CD30 IHCs. Detected mutations in paired regions are displayed to infer clonal relatedness between the CLL/SLL and CHL components.

### Patients with type 1 and type 2 CHL-RT show similar demographics and clinical outcomes

Patients with either type 1 or type 2 CHL-RT exhibited similar gender distributions, with a male predominance in both groups (77.8% in type 1 vs. 72% in type 2, p=0.79). Additionally, both groups demonstrated comparable ages at the time of CHL-RT diagnosis (median: 71 vs. 70.5 years for type 1 vs. type 2, p=0.14) (**Figure 6A**). The interval between CLL/SLL and CHL diagnosis was also similar, with median times of 5.3 years for type 1 and 3.5 years for type 2 CHL-RT (p=0.16) (**Figure 6B**). Finally, overall survival did not differ significantly between the two groups, whether measured from the time of CLL/SLL diagnosis (median survival: 10.5 vs. 15.0 years for type 1 vs. type 2, p=0.95) (**Figure 6C**) or from the time of CHL diagnosis (median survival: 2.7 vs. 5.3 years for type 1 vs. type 2, p=0.12) (**Figure 6D**).

**Figure 6.**
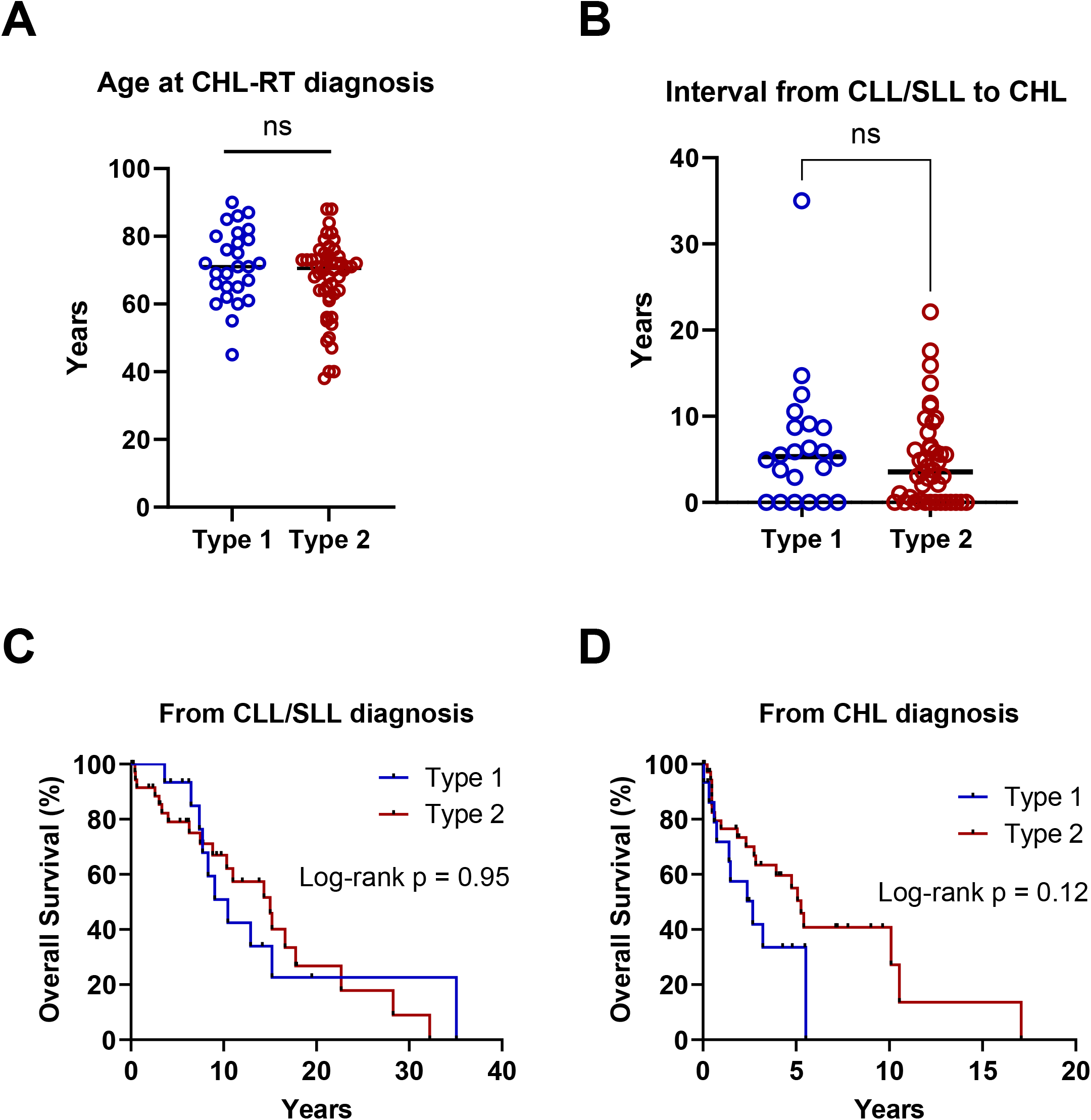
Clinical characteristics and outcomes of type 1 vs type 2 CHL□RT. (A) Age at CHL-RT diagnosis for type 1 vs type 2 patients (median values shown). Black bars indicate group median. ns, statistically not significant. (B) Interval from CLL/SLL to CHL-RT diagnosis in type 1 vs type 2 CHL□ RT. Black bars indicate group median. ns, statistically not significant. (C) Kaplan-Meier overall survival (OS) from time of CLL/SLL diagnosis for type 1 vs type 2 CHL□ RT. (D) Kaplan-Meier OS from time of CHL-RT diagnosis for type 1 vs type 2 CHL□ RT.

## DISCUSSION

We provide novel evidence that HRS cells in CHL-RT and *de novo* CHL share a common immune evasion phenotype, including frequent *PD-L1/PD-L2* gain/amplification, establishing for the first time a shared biological and genetic basis between these two conditions.

A key distinction, however, emerged in the tumor microenvironment. PD-L1 expression on HRS cells was significantly lower in type 1 CHL-RT as compared to both type 2 CHL-RT and *de novo* CHL. Using mIF, we demonstrated that this difference reflects a fundamentally less immunosuppressive TME in type 1 CHL-RT, with lower number of both CTLA-4□ and PD-1□ T cells. In contrast, these immunosuppressive T cell populations were comparably dense in type 2 CHL-RT and *de novo* CHL. Collectively, these findings indicate that type 2 CHL-RT fully recapitulates the immunosuppressive microenvironment of *de novo* CHL, while type 1 CHL-RT represents an earlier stage of incomplete microenvironmental reprogramming. This interpretation is further supported by the direct observation (including our cohort) of patients who demonstrated type 1 histology on initial biopsy with subsequent progression to type 2 CHL-RT (6,9), suggesting a chronological evolution from type 1 to type 2. This shared suppressive TME provides a biological rationale for exploring immune checkpoint inhibitors in CHL-RT, particularly in type 2 disease, where the microenvironment most closely mirrors that of de novo CHL, a setting in which PD-1 blockade has demonstrated established clinical efficacy(24). The less pronounced but present immune-evasive phenotype in type 1 CHL-RT similarly warrants investigation into whether checkpoint inhibition may have a role in this subtype.

Our NGS analysis reveals a distinct mutational landscape that identifies potential genetic risk factors for CHL-RT. Most notably, *XPO1* hotspot mutations (p.E571) were identified in 16.2% of CLL/SLL with CHL-RT, comparable to the 13.1% frequency reported in flow sorted HRS cells from *de novo* CHL(19,20), suggesting that *XPO1* mutations potentially represent a shared genetic risk factor for Hodgkin lymphomagenesis in both disease contexts. This finding is therapeutically relevant, as XPO1 is targetable by the small-molecule inhibitor selinexor (KPT-330), which has demonstrated preclinical efficacy in lymphoma and leukemia models and is now in clinical use. Interestingly, *XPO1* mutations were more frequently observed in CLL/SLL cases with type 1 CHL-RT (27.8%) compared with those with type 2 CHL-RT (5.3%); however, this difference did not reach statistical significance (p = 0.09), likely due to the relatively small sample size. It remains unclear whether CLL/SLL cases associated with type 1 versus type 2 CHL-RT harbor fundamentally distinct mutational profiles, or whether differences in HRS cell biology between the two subtypes account for the observed variability. Larger-scale studies with improved single cell sequencing technology will be required to validate these observations and to further elucidate the biological differences between these entities. Beyond *XPO1*, enrichment of mutations in *FBXW7* (regulating c-Myc/Notch turnover), NF-κB pathway genes (*BIRC3, TRAF3*), and immune-related genes (*HLA-A*) suggests that enhanced NF-κB signaling and acquisition of immune evasion, hallmarks of *de novo* CHL, may contribute to CHL-RT predisposition.

Despite these enrichments, the overall genetic profile of CLL/SLL with CHL-RT remains broadly similar to conventional CLL/SLL. Highly recurrent *de novo* CHL mutations (*B2M, SOCS1, STAT6, TNFAIP3, ITPKB*) were not detected, and no individual gene exceeded a 20% mutation frequency, suggesting that CHL-RT risk reflects complex polygenic interactions rather than single driver events. Notably, the genetic risk profile of CHL-RT appears distinct from DLBCL-RT, which is characterized by *TP53* disruption(15-17), *CDKN2A/B* inactivation(16,18), *NOTCH1* mutations(16,17), and *MYC* activation(15-17), none of which were enriched in our cohort (copy number data were unavailable to assess *CDKN2A/B*). This distinction underscores that CHL-RT and DLBCL-RT, while both arising from CLL/SLL, are driven by fundamentally different genetic programs.

This study has several limitations. Its retrospective design resulted in incomplete treatment histories for many patients. NGS was performed on bulk CLL/SLL tissue rather than isolated HRS cells, precluding definitive conclusions about HRS-specific mutations. Furthermore, clonal relationship analyses between HRS and CLL/SLL cells were feasible in only a limited subset of cases. Additionally, the absence of copy number alteration data other than *PD-L1/PD-L2* limits direct comparison with *de novo* CHL. Future studies employing single-cell sequencing or laser-capture microdissection of HRS cells will be essential to fully characterize the molecular underpinnings of CHL-RT and to determine whether the mutations identified in bulk CLL/SLL tissue are also present in the HRS compartment.

In summary, this study provides the first integrated immunophenotypic and genomic characterization of CHL-RT, establishing biological links to *de novo* CHL. These findings lay a foundation for future mechanistic and therapeutic investigations of this rare and biologically distinct form of Richter transformation.

## METHODS

### Patient cohort

A total of 77 CLL/SLL patients with CHL-RT were identified and collected from 3 academic institutions in the United States (27 from Memorial Sloan Kettering Cancer Center (MSKCC), 28 from Mayo Clinic, and 22 from National Cancer Institute). A portion of these patients have been previously reported in separate studies(6,9). CHL□RT diagnoses were identified over a study period extending from 1995 to 2022 (8 cases were diagnosed between 1995 and 2004, 30 cases between 2005 and 2014, and 39 cases between 2015 and 2025). This cohort included 27 type 1 and 50 type 2 patients, with ten of the type 2 patients exhibiting pathology samples that showed only CHL without morphological and immunophenotypic evidence of previously diagnosed CLL/SLL. Notably, four patients demonstrated both type 1 and type 2 diseases in separate pathological samples; for the purposes of this study, these patients were classified as having type 2 diseases. The diagnosis was rendered and reviewed by one of the 3 pathologists (ESJ, RK, and WX) and confirmed by a central review (YM and WX). Detailed information on patient numbers from each contributing institution is shown in **Figure 1A**. Patient demographics, HRS cell percentages within CLL/SLL tissue, testing modalities, and clinical features are summarized in **Supplementary Table 1**. A cohort of 48 *de novo* CHL cases diagnosed with 87.5% nodular sclerosis subtype at MSKCC was also included for comparison (25). The study was conducted under IRB-approved protocols at each of the contributing institutions.

### Immunohistochemistry (IHC) and in situ hybridization (ISH)

IHC and ISH staining were performed on 5-micron formalin-fixed paraffin-embedded (FFPE) tissue slides for all cases with sufficient tissue samples and HRS cells available for evaluation. The following antibodies or probe were used: PD-L1 (clone: E1L3N), PD-L2 (clone: D7U8C), MHC-I (clone: A4), B2M (clone: A0072), HLA-DR (MHC-II) (clone: TAL 1B5), CIITA (clone: 7-1H), EBV-encoded RNA (EBER) ISH (Ventana, 760-1209). Appropriate positive and negative controls were included and confirmed for all antibodies. A 30% cut-off for IHC was applied to assess the expression of PD-L1, PD-L2, MHC-I, B2M, MHC-II, and CIITA in HRS cells. Expression was classified as positive if more than 30% of the HRS cells showed staining of the IHC markers.

### Multiplexed immunofluorescence (mIF)

Tissue sections were stained using a six-marker multiplex immunofluorescence (mIF) assay with fluorophore-conjugated antibodies targeting the specific antigens: CD68 (Opal 480), PAX% (Opal 520), CD86 (Opal 570), LEF1 (Opal 620), CTLA-4 (Opal 690), and PD-1 (Opal 780). Staining was performed on the Leica BOND RX automated platform (Leica Biosystems). Heat-induced epitope retrieval (HIER) was conducted using Epitope Retrieval Solution 2 (ER2, Leica) for 30 minutes. Each primary antibody—CD68 (clone PM-G1, Agilent Technologies), PAX5 (clone D7H5X, Cell Signaling Technology), CD86 (clone E2G8P, Cell Signaling Technology), CTLA-4 (clone CAL49, abcam), LEF1 (clone EPR2029Y, abcam), and PD-1 (clone NAT105, Millipore Sigma) —was applied sequentially for 30 minutes at room temperature. Antibody dilutions were optimized through prior titration on normal and neoplastic control tissues. Secondary detection employed species-specific SuperBoost™ Poly HRP antibodies (ThermoFisher), followed by tyramide signal amplification using the Opal 6-Plex Detection Kit (Akoya Biosciences). After each round, antibodies were stripped via HIER with ER2. Slides were counterstained with 4′,6-diamidino-2-phenylindole (DAPI) and mounted using ProLong™ Gold antifade reagent (ThermoFisher).

mIF-stained slides were scanned at 20× magnification using a PhenoImager (Akoya/Quanterix) whole-slide scanner. Whole-slide images were imported into the HALO image analysis platform (version (4.0), Indica Lab). Cell segmentation was performed using the HALO (Nuclei Seg V2) module, with nuclei identified based on DAPI staining. Marker positivity thresholds were established using internal positive and negative controls and applied uniformly across all samples. Subsequently, 150 × 150 µm regions of interest (ROIs) centered on Hodgkin and Reed-Sternberg (HRS) cells were manually annotated to evaluate the tumor microenvironment within a 75 µm radius. HRS cells were identified based on nuclear size and morphology, as well as expression of CD86(14). Quantitative outputs, including the number of PD-1□ T cells (defined by co-expression of LEF1 and PD-1 in the absence of all other markers), CTLA-4□ T cells (defined by co-expression of LEF1 and CTLA-4 in the absence of all other markers), B cells (defined by expression of PAX5), and histiocytes (defined by expression of CD68) within ROIs, were exported for downstream statistical analysis. A total of 25 cells from three type 1 CHL-RT cases, 25 cells from three type 2 CHL-RT cases, and 17 cells from two *de novo* CHL cases were analyzed.

### Flow cytometric analysis

Flow cytometric studies were performed on all samples in a standard manner as part of routine clinical practice at MSKCC. Tissue biopsy specimens were finely minced in RPMI 1640 growth medium using scalpel blades, then filtered and resuspended in RPMI. Needle aspirates and biopsy needle rinses were suspended in RPMI. Cells in RPMI were then stained with a 21-antibody assay (CD2, CD3, CD4, CD5, CD7, CD8, CD10, CD14, CD19, CD20, CD22, CD25, CD26, CD38, CD45, CD56, CD279 (PD-1), TCRγδ, TCRCβ1, Kappa, Lambda) designed to evaluate both B and T cells(26). Due to availability and periodic changes in clinical laboratory protocols, some cases were stained with only portions of this modular panel separately. Following staining and incubation, cells were lysed, fixed, washed, and resuspended. Up to 500,000 cells were acquired on a Fluorescence-activated cell sorting (FACS) Canto 10 color flow cytometer (BD), BD LSR Fortessa X-20 20 color flow cytometer (BD), or BD FACSymphony A3 28 color flow cytometer (BD). Results were analyzed with custom Woodlist software (generous gift of Wood BL, Children’s Hospital of Los Angeles, Los Angeles, CA, USA). All assays were initially assessed by a single pathologist (A.C.).

### Combined CD30 immunofluorescence and *PD-L1/PD-L2* FISH analysis

Following immunophenotyping with mouse anti-CD30 antibody (clone: Ber-H2) labeled with aminomethylcoumarin acetate (AMCA), a blue fluorescent dye, the tissue slides with coverslips were briefly reviewed and documented under a fluorescence microscope to assess the intensity and quality of CD30 staining. After removing the coverslips, the slides were washed in 2× saline-sodium citrate (SSC) buffer at room temperature for 5 minutes, then fixed in 10% neutral buffered formalin for 20 minutes. The slides were subsequently dehydrated in ascending alcohol concentrations (70%, 85%, and 100%) for 5 minutes each. Next, the slides underwent pretreatment with a 20 mM citrate buffer and 1% NP-40 mixture (pH 6.0-6.5) for 10 minutes, followed by protease digestion at 40°C for 10-15 minutes (Abbott Molecular, Des Plaines, IL). The dehydration process was repeated with 2-minute immersions in 70%, 85%, and 100% alcohol. For fluorescence in situ hybridization (FISH), probes for *PD-L1* (Spectrum Orange) and *PD-L2* (Spectrum Green) (both from Empire Genomics, Buffalo, NY) were used, along with *CEP9* (Spectrum Aqua) (Abbott Molecular, Des Plaine, IL) as an internal control for chromosome 9 copy number (all from Genomic Empire, Buffalo, NY). FISH testing follows the standard protocol. Briefly, after applying the FISH probes to the tissue areas, both tissue and probes were co-denatured at 94º C for 7 minutes, and then incubated at 37º C overnight, followed by post-hybridization washing in 2xSSC/0.3% NP-40 at 77º C for one minute. The sections were counterstained with an antifade medium lacking DAPI (Vector Laboratories, Burlingame, CA). Slides were examined under a fluorescence microscope equipped with appropriate filters for CD30 immunofluorescence and *PD-L1/PD-L2* and *CEP9* probes. Signal analysis was conducted in conjunction with tissue morphology, focusing on areas with strong CD30+ cells. Probe copy numbers were counted in up to 50 cells per case, where possible. The following criteria were used for interpreting *PD-L1/PD-L2* FISH results according to previous publication(11): amplification: *PD-L1/PD-L2/CEP9* ratio > 3.0; gain: *PD-L1/PD-L2/CEP9* ratio > 1.0 but less than <3.0; polysomy: *PD-L1/PD-L2/CEP9* ratio ≈ 1.0 but more than two copies for each probe.

### Next-generation sequencing (NGS)

The targeted NGS analysis was conducted using our laboratory-developed MSK-IMPACT (Integrated Mutation Profiling of Actionable Cancer Targets)-Heme assay, a hybrid-capture-based platform designed to capture all canonical exons of 400 key oncogenes and tumor suppressor genes implicated in hematologic malignancies(27). Gene list is available in **Supplementary Table 2**. This includes genes that are either druggable by approved therapies or are targets of experimental therapies currently under investigation in clinical trials at our institution. Unlike the paired sequencing approach used in routine clinical practice, which incorporates matched normal tissue to distinguish somatic from germline variants, the NGS analyses in this study were performed without matched normal controls. Consequently, germline variants and patient-specific single nucleotide polymorphisms (SNPs) may be present in the data. To minimize the potential bias introduced by such events, we focused exclusively on pathogenic or likely pathogenic variants for downstream analysis. These variants were identified using several databases, including ClinVar(28), OncoKB(29), and COSMIC (Catalogue Of Somatic Mutations In Cancer)(30).

NGS was performed on bulk CLL/SLL FFPE tissue from 17 patients with type 1 CHL-RT and 15 patients with type 2 CHL-RT, two separate bulk CLL/SLL FFPE tissue from a patient who exhibited either type 1 or type 2 CHL-RT in different samples, and bulk FFPE tissue from three patients with type 2 CHL-RT with samples showing only CHL with a high density of HRS cells (comprising more than 10% cellularity) but no morphologic and flow cytometric evidence of previously diagnosed CLL/SLL. In addition, macrodissection was performed on samples from three patients with type 2 CHL-RT who displayed distinct areas that were enriched for either CLL/SLL or CHL, allowing for separate analysis of the CLL/SLL and CHL components. In total, NGS was performed on 43 samples from 39 patients, comprising 18 CLL/SLL samples from type 1 CHL-RT, 19 CLL/SLL samples from type 2 CHL-RT, and 6 CHL-enriched samples from type 2 CHL-RT. It should be noted that bulk CLL/SLL sequencing inherently included rare HRS cells, which constitute less than 5% of total cellularity.

### Statistical analysis

Numerical data were analyzed using Student’s t-test. Categorical data were compared by Fisher’s exact test. Overall survival was calculated from the initial date of diagnosis until the date of death or last follow-up. Survival data were visualized using Kaplan-Meier curves and analyzed with the two-sided log-rank (Mantel-Cox) test. The statistical analysis was performed using GraphPad Prism version 10.0.0 software (San Diego, California; www.graphpad.com) or R (version 4.2.2). The oncoplot was generated by ggplot2 in R (version 4.2.2). Statistical significance was defined as a p-value less than 0.05.

## Data Availability

Original data can be shared upon request by contacting the corresponding author.

## Acknowledgment

We would like to thank Ms. Irina Linkov (Core facility, MSKCC) for providing technical support in performing immunohistochemical study.

## FIGURE LEGENDS

**Table 1:** Summary of patient cohort.

|  |  | **CHL-RT type** |  |
| --- | --- | --- | --- |
|  |  | **Type 1**<br>**(n=27)** | **Type 2**<br>**(n=50)** |
| **Sex,**<br>**male (%)** |  | 21 (77.8%) | 36 (72%) |
| **Age at CLL/SLL diagnosis,**<br>**median (range)** |  | 63 (45 to 86) | 64 (37 to 87) |
| **Age at CHL-RT diagnosis,**<br>**median (range)** |  | 71 (45 to 90) | 70.5 (38 to 88) |
| **HRS cells % in CLL/SLL tissue,**<br>**number (%)** | ≤ 5% | 26 (96.3%) | 37 (74%) |
|  | 5-10% | 0 | 6 (12%) |
|  | ≥ 10% | 1 (3.7%) | 7 (14%) |
| **Years from CLL/SLL to CHL-RT diagnosis,**<br>**median (range)** |  | 5.5 (0 to 35.0) | 3.5 (0 to 22.1) |
| **IHC for immune markers tested,**<br>**number (%)** |  | 18 (66.7%) | 31 (60.0%) |
| **Bulk NGS for CLL/SLL tested,**<br>**number (%)** |  | 18 (66.7%) | 20 (40.0%) |
| **FISH for *PD-L1*/*PD-L2* tested,**<br>**number (%)** |  | 4 (14.8%) | 8 (16.0%) |
| **Multiplex IF tested,**<br>**number (%)** |  | 3 (11.1%) | 3 (6.0%) |

