## supplemental table 1 and table 2 for "Genetic risk and immune dysregulation of classic Hodgkin lymphoma transformation of chronic lymphocytic leukemia/small lymphocytic lymphoma: a multicentric study"

### Supplementary table 1:

#### Patient demographics, clinicopathologic characteristics, and ancillary testing

| | | Demographics | | | | | Ancillary tests | | | IHC and ISH results | | | | | | | | | Treatments and outcomes | | |
|---|---|---|---|---|---|---|---|---|---|---|---|---|---|---|---|---|---|---|---|---|---|
| **ID** | **CLL/SLL type** | **Sex** | **Age range at CLL/SLL dx** | **Age range at CHL-RT dx** | **HRS cell % in CLL/SLL** | **Months between CLL/SLL and CHL** | **NGS tested** | **FISH tested** | **mIF tested** | **EBER** | **CD30** | **CD15** | **PD-L1** | **PD-L2** | **MHC-I** | **B2M** | **MHC-II** | **CIITA** | **CLL treatment** | **CHL treatment** | **Outcome** |
| **1** | Type 1 | M | 76-80 | 76-80 | <5% | 0 | Yes | No | No | - | + | + | - | - | + | - | - | + | NA | NA | Alive |
| **2** | Type 1 | F | 61-65 | 76-80 | <5% | 176.4 | Yes | No | Yes | + | + | - | NA | NA | NA | + | NA | + | PCR >> ibrutinib | AVD + ibrutinib >> V + OB | Alive |
| **3** | Type 1 | M | 71-75 | 76-80 | <5% | 70.5 | No | No | No | - | + | + | NA | NA | NA | NA | NA | NA | NA | R-CHOP >> R + benda >> ibrutinib >> R-GCVP >> palliative RT | Deceased |
| **4** | Type 1 | F | 81-85 | 86-90 | ~5% | 66.3 | No | No | No | NA | + | - | NA | NA | NA | NA | NA | NA | ibrutinib >> observation | NA | Alive |
| **5** | Type 1 | M | NA | 71-75 | <5% | NA | No | No | No | - | + | + | NA | NA | NA | NA | NA | NA | NA | NA | Alive |
| **6** | Type 1 | M | NA | 86-90 | <5% | NA | No | No | No | - | + | - | NA | NA | NA | NA | NA | NA | NA | NA | Alive |
| **7** | Type 1 | M | 41-45 | 41-45 | <5% | 0 | No | No | No | NA | + | + | NA | NA | NA | NA | NA | NA | NA | ABVD | Alive |
| **8** | Type 1 | M | 56-60 | 56-60 | <5% | 0 | Yes | No | No | + | + | + | + | - | NA | - | - | + | NA | NA | NA |
| **9** | Type 1 | F | 86-90 | 86-90 | <5% | 0 | Yes | No | No | - | + | + | - | NA | NA | NA | - | NA | NA | NA | NA |
| **10** | Type 1 | M | NA | 51-55 | ~5% | NA | Yes | No | No | + | + | + | + | - | + | + | - | + | NA | NA | NA |
| **11** | Type 1 | M | 56-60 | 61-65 | <5% | 109.6 | Yes | No | No | + | + | - | - | - | NA | - | NA | + | NA | NA | NA |
| **12** | Type 1 | M | 56-60 | 56-60 | <5% | 48.7 | Yes | No | No | + | + | + | + | - | + | + | + | - | NA | NA | NA |
| **13** | Type 1 | M | 66-70 | 66-70 | <5% | 0 | Yes | No | Yes | + | + | - | - | - | + | - | + | + | NA | NA | NA |
| **14** | Type 1 | F | NA | 66-70 | ~5% | NA | Yes | Yes | No | - | + | + | - | - | + | + | + | + | NA | NA | NA |
| **15** | Type 1 | M | NA | 61-65 | <5% | NA | Yes | No | No | - | + | + | + | - | + | + | + | + | NA | NA | NA |
| **16** | Type 1 | M | NA | 61-65 | 10-20% | NA | Yes | No | No | - | + | + | - | NA | NA | + | + | NA | NA | NA | NA |
| **17** | Type 1 | F | 66-70 | 71-75 | <5% | 104.3 | No | No | No | + | + | - | NA | NA | NA | NA | NA | NA | NA | NA | Alive |
| **18** | Type 1 | M | 56-60 | 66-70 | <5% | 70.7 | No | No | No | + | + | - | NA | NA | NA | NA | NA | NA | NA | NA | Deceased |
| **19** | Type 1 | F | 61-65 | 66-70 | <5% | 59.2 | Yes | No | No | + | + | + | + | - | + | + | - | + | NA | NA | Deceased |
| **20** | Type 1 | M | 66-70 | 71-75 | ~5% | 150.3 | Yes | Yes | Yes | + | + | - | - | - | + | - | + | + | NA | NA | Deceased |
| **21** | Type 1 | M | 66-70 | 71-75 | <5% | 75.7 | Yes | No | No | + | + | + | + | - | - | + | + | + | NA | NA | Deceased |
| **22** | Type 1 | M | 81-85 | 81-85 | <5% | 34.8 | Yes | No | No | + | + | - | NA | NA | NA | NA | NA | NA | NA | NA | Deceased |
| **23** | Type 1 | M | 46-50 | 81-85 | <5% | 420.4 | No | No | No | + | + | - | NA | NA | NA | NA | NA | NA | NA | NA | Deceased |
| **24** | Type 1 | M | 71-75 | 81-85 | <5% | 126.4 | Yes | No | No | - | + | + | NA | NA | NA | NA | NA | NA | NA | NA | Deceased |
| **25** | Type 1 | M | 61-65 | 71-75 | <5% | 104.5 | Yes | Yes | No | NA | + | + | + | - | - | + | + | + | NA | NA | Deceased |
| **26** | Type 1 | M | 56-60 | 61-65 | <5% | 61.4 | Yes | Yes | No | + | + | + | + | - | + | - | - | + | NA | NA | Deceased |
| **27** | Type 1 | M | 71-75 | 76-80 | <5% | 45.2 | No | | No | + | + | - | NA | NA | NA | NA | NA | NA | NA | NA | Alive |
| **28** | Type 2 | M | 41-45 | 46-50 | ~10% | 58.8 | Yes | Yes | No | - | + | + | NA | NA | NA | NA | NA | + | PCR >> CAR-T >> RT >> ibrutinib | ABVD >> R + idelalisib >> GCV >> OB >> benda + OB >> SCT >> pembrolizumab | Alive |

| | | | | | | | | | | | | | | | | | | | | | |
|---|---|---|---|---|---|---|---|---|---|---|---|---|---|---|---|---|---|---|---|---|---|
| **29 #** | Type 2 | M | 61-65 | 61-65 | <5% | 0 | Yes | No | No | + | + | - | NA | NA | NA | NA | NA | + | NA | R-EPOCH + V >> SCT | Alive |
| **30 *** | Type 2 | M | 76-80 | 76-80 | ~5% | 0.25 | Yes | Yes | No | - | + | - | + | NA | NA | + | - | + | NA | NA | Alive |
| **31** | Type 2 | M | 61-65 | 76-80 | <5% | 190.9 | No | No | No | + | + | + | NA | NA | NA | NA | NA | NA | NA | NA | Deceased |
| **32** | Type 2 | M | 61-65 | 61-65 | 5-10% | 0 | No | No | No | NA | + | + | NA | NA | NA | NA | NA | NA | NA | ABVD >> AVD | Alive |
| **33** | Type 2 | M | 46-50 | 46-50 | 5-10% | 0 | No | No | No | + | + | - | + | - | + | - | + | + | NA | NA | Deceased |
| **34** | Type 2 | M | 66-70 | 66-70 | <5% | 50.3 | No | No | No | NA | + | + | + | - | + | + | - | + | acalabrutinib | BV-AVD >> nivolumab + AD >> OB + V | Alive |
| **35** | Type 2 | M | 61-65 | 61-65 | 5-10% | 0 | No | No | No | NA | + | + | NA | NA | NA | NA | NA | NA | NA | BV >> AVD >> BV | Alive |
| **36** | Type 2 | M | 71-75 | 76-80 | <5% | 66.5 | Yes | No | No | NA | + | - | + | - | + | + | + | + | observation | BV >> AVD | Alive |
| **37** | Type 2 | M | 71-75 | 71-75 | <5% | 0 | Yes | No | Yes | - | + | - | NA | NA | NA | NA | NA | + | NA | NA | Deceased |
| **38** | Type 2 | F | 61-65 | 61-65 | <5% | 0 | No | No | No | + | + | + | NA | NA | NA | NA | NA | NA | NA | NA | Alive |
| **39** | Type 2 | M | 51-55 | 56-60 | <5% | 45.1 | No | No | No | NA | + | - | + | - | + | NA | + | + | ibrutinib >> R | BV + AVD | Alive |
| **40** | Type 2 | M | 36-40 | 36-40 | <5% | 0 | No | No | No | + | + | + | NA | NA | NA | NA | NA | NA | NA | ABVD >> RT | Alive |
| **41** | Type 2 | F | 61-65 | 66-70 | <5% | 56.7 | No | No | No | NA | + | + | NA | NA | NA | NA | NA | NA | observation >> FCR >> BR >> ibrutinib | ABVD | Deceased |
| **42** | Type 2 | M | 86-90 | 86-90 | <5% | 12.1 | No | No | No | - | + | + | NA | NA | NA | NA | NA | NA | NA | COPP >> BV >> ibrutinib | Deceased |
| **43** | Type 2 | M | 36-40 | 36-40 | <5% | 6.8 | No | No | No | + | + | - | NA | NA | NA | NA | NA | NA | FCR | ABVD | Alive |
| **44** | Type 2 | M | 56-60 | 81-85 | <5% | 265.4 | No | No | No | NA | + | + | NA | NA | NA | NA | NA | NA | observation >> bleomycin + CHOP >> R | R-CEPP >> BV >> acalabrutinib | Deceased |
| **45** | Type 2 | M | 61-65 | 66-70 | <5% | 73 | No | No | No | NA | + | + | NA | NA | NA | NA | NA | NA | observation | ABVD >> GVD >> SCT >> ibrutinib >> V | Deceased |
| **46** | Type 2 | F | NA | 36-40 | 5-10% | NA | No | No | No | NA | + | + | NA | NA | NA | NA | NA | NA | NA | NA | Alive |
| **47** | Type 2 | M | 71-75 | 71-75 | <5% | 2.8 | No | No | No | - | + | - | NA | NA | NA | NA | NA | NA | NA | R-CHOP >> RT | Alive |
| **48** | Type 2 | M | 66-70 | 71-75 | <5% | 36.5 | No | No | No | NA | + | - | NA | NA | NA | NA | NA | NA | V + OB | BV >> AVD >> BV | Alive |
| **49** | Type 2 | F | 56-60 | 71-75 | 5-10% | 166.3 | No | No | No | + | + | + | NA | NA | NA | NA | NA | NA | observation >> ibrutinib >> V | benda + ibrutinib >> AVD >> BV | Alive |
| **50** | Type 2 | M | 66-70 | 71-75 | ~5% | 36.5 | Yes | No | No | - | + | + | + | - | NA | + | - | NA | NA | NA | NA |
| **51** | Type 2 | F | 66-70 | 66-70 | ~10% | 36.5 | Yes | Yes | No | - | + | + | + | - | + | - | - | + | NA | NA | NA |
| **52** | Type 2 | M | 66-70 | 66-70 | <5% | 0 | Yes | No | No | + | + | + | + | - | + | + | + | - | NA | NA | NA |
| **53** | Type 2 | M | 71-75 | 71-75 | ~10% | 0 | Yes | Yes | No | - | + | + | + | - | - | - | - | + | NA | NA | NA |
| **54** | Type 2 | M | NA | 66-70 | <5% | NA | Yes | No | No | + | + | + | + | NA | NA | + | - | + | NA | NA | NA |
| **55** | Type 2 | F | NA | 81-85 | 10-20% | NA | Yes | Yes | Yes | + | + | + | + | - | - | + | - | + | NA | NA | NA |
| **56** | Type 2 | M | 66-70 | 76-80 | <5% | 97.4 | No | No | No | - | + | + | + | - | + | + | - | + | NA | NA | NA |
| **57** | Type 2 | F | NA | 61-65 | <5% | NA | No | No | No | NA | + | + | + | - | - | - | - | + | NA | NA | NA |
| **58** | Type 2 | M | 66-70 | 71-75 | <5% | 24.4 | Yes | No | No | + | + | + | + | - | - | NA | - | + | NA | NA | NA |
| **59** | Type 2 | F | NA | 71-75 | 20-30% | NA | Yes | Yes | No | + | + | + | + | NA | NA | - | + | NA | NA | NA | NA |
| **60** | Type 2 | F | 46-50 | 46-50 | <5% | 0 | No | No | No | + | + | + | + | - | + | - | - | + | NA | NA | NA |
| **61** | Type 2 | F | NA | 56-60 | <5% | NA | No | No | No | + | + | - | + | NA | NA | - | + | + | NA | NA | NA |
| **62** | Type 2 | M | NA | 71-75 | ~5% | NA | Yes | No | No | + | + | - | + | - | + | - | + | + | NA | NA | NA |
| **63 *** | Type 2 | M | 56-60 | 66-70 | <5% | 66.8 | Yes | Yes | No | + | + | + | + | NA | NA | - | + | - | NA | NA | Alive |
| **64** | Type 2 | M | 46-50 | 51-55 | <5% | 58.5 | Yes | No | No | + | + | + | + | - | + | + | - | + | NA | NA | Deceased |
| **65** | Type 2 | F | 66-70 | 71-75 | ~10% | 25.2 | Yes | Yes | Yes | NA | + | + | + | - | + | + | + | + | NA | NA | Deceased |

| | | | | | | | | | | | | | | | | | | | | | |
|---|---|---|---|---|---|---|---|---|---|---|---|---|---|---|---|---|---|---|---|---|---|
| **66** | Type 2 | F | 61-65 | 71-75 | <5% | 134.2 | No | No | No | + | + | + | NA | NA | NA | NA | NA | NA | NA | NA | Deceased |
| **67** | Type 2 | M | 61-65 | 61-65 | <5% | 0.1 | No | No | No | + | + | - | NA | NA | NA | NA | NA | NA | NA | NA | Deceased |
| **68** | Type 2 | M | 56-60 | 66-70 | 10-20% | 117.5 | Yes | No | No | + | + | + | + | + | - | - | + | + | NA | NA | Deceased |
| **69** | Type 2 | M | 61-65 | 71-75 | <5% | 112.2 | Yes | No | No | - | + | + | + | + | + | + | + | + | NA | NA | Deceased |
| **70** | Type 2 | M | 71-75 | 81-85 | <5% | 117 | No | No | No | + | + | + | NA | NA | NA | NA | NA | NA | NA | NA | Deceased |
| **71 *** | Type 2 | M | 71-75 | 76-80 | <5% | 42.4 | Yes | No | No | - | + | - | + | NA | - | + | + | + | NA | NA | Deceased |
| **72** | Type 2 | M | 41-45 | 51-55 | <5% | 138.2 | No | No | No | + | + | + | NA | NA | NA | NA | NA | NA | NA | NA | Deceased |
| **73** | Type 2 | F | 66-70 | 71-75 | <5% | 79.1 | No | No | No | - | + | + | NA | NA | NA | NA | NA | NA | NA | NA | Alive |
| **74** | Type 2 | M | 56-60 | 61-65 | ~5% | 69.7 | Yes | No | No | + | + | + | + | NA | NA | + | - | + | NA | NA | Deceased |
| **75** | Type 2 | M | 61-65 | 71-75 | <5% | 74.9 | No | No | No | - | + | + | NA | NA | NA | NA | NA | NA | NA | NA | Deceased |
| **76** | Type 2 | M | 66-70 | 71-75 | 5-10% | 33.9 | No | No | No | - | + | + | + | - | + | + | + | + | NA | NA | Deceased |
| **77** | Type 2 | F | 66-70 | 86-90 | <5% | 211 | No | No | No | + | + | - | + | - | + | + | + | + | NA | NA | Deceased |

Abbreviations: **ABVD**: doxorubicin, bleomycin, vinblastine, dacarbazine. **AD**: doxorubicin and dacarbazine. **AVD**: doxorubicin, vinblastine, dacarbazine. **Benda**: bendamustine. **BR**: bendamustine and rituximab. **BV**: brentuximab vedotin. **CAR-T**: chimeric antigen receptor-T cell therapy. **CEPP**: cyclophosphamide, etoposide, procarbazine, and prednisone. **CHOP**: cyclophosphamide, doxorubicin, vincristine, and prednisone. **COPP**: cyclophosphamide, vincristine, procarbazine hydrochloride, and prednisone. **EPOCH**: etoposide phosphate, prednisone, vincristine, cyclophosphamide, and doxorubicin. **FCR**: fludarabine, cyclophosphamide, and rituximab. **GCV**: gemcitabine, cyclophosphamide, vincristine. **GCVP**: gemcitabine, cyclophosphamide, vincristine, prednisolone. **GVD**: gemcitabine, vincristine, and doxorubicin. **IF**: immunofluorescence. **IHC**: Immunohistochemistry. **ISH**: In-situ hybridization. **mIF**: multiplex immunofluorescence. **OB**: obinutuzumab. **PCR**: pentostatin, cyclophosphamide, and rituximab. **R**: rituximab. **RT**: radiation therapy. **SCT**: stem cell transplantation. **V**: venetoclax.

Note: # For this patient, two samples—one with type 1 and another with type 2 CLL/SLL—were submitted for NGS studies.

* (NGS) studies were conducted on macrodissected CLL/SLL-enriched and CHL-enriched areas separately for each of these patients.

#### Supplementary table 2:

#### Gene list for MSK IMPACT-Heme

| | | | | | | | | | |
|---|---|---|---|---|---|---|---|---|---|
| *ABL1* | *ACTG1* | *AKT1* | *AKT2* | *AKT3* | *ALK* | *ALOX12B* | *AMER1* | *APC* | *AR* |
| *ARAF* | *ARHGEF28* | *ARID1A* | *ARID1B* | *ARID2* | *ARID3A* | *ARID3B* | *ARID3C* | *ARID4A* | *ARID4B* |
| *ARID5A* | *ARID5B* | *ASXL1* | *ASXL2* | *ATM* | *ATP6AP1* | *ATP6V1B2* | *ATR* | *ATRX* | *ATXN2* |
| *AURKA* | *AURKB* | *AXIN1* | *AXL* | *B2M* | *BACH2* | *BAP1* | *BARD1* | *BCL10* | *BCL11B* |
| *BCL2* | *BCL6* | *BCOR* | *BCORL1* | *BCR* | *BIRC3* | *BLM* | *BRAF* | *BRCA1* | *BRCA2* |
| *BRD4* | *BRIP1* | *BTG1* | *BTK* | *CALR* | *CARD11* | *CASP8* | *CBFB* | *CBL* | *CCND1* |
| *CCND2* | *CCND3* | *CCNE1* | *CD274* | *CD28* | *CD58* | *CD79A* | *CD79B* | *CDC73* | *CDH1* |
| *CDK12* | *CDK4* | *CDK6* | *CDK8* | *CDKN1B* | *CDKN2A* | *CDKN2B* | *CDKN2C* | *CEBPA* | *CHEK1* |
| *CHEK2* | *CIC* | *CIITA* | *CRBN* | *CREBBP* | *CRKL* | *CRLF2* | *CSF1R* | *CSF3R* | *CTCF* |
| *CTNNB1* | *CUX1* | *CXCR4* | *CYLD* | *DAXX* | *DDR2* | *DDX3X* | *DIS3* | *DNMT3A* | *DOT1L* |
| *DTX1* | *DUSP22* | *EED* | *EGFR* | *EGR1* | *EP300* | *EP400* | *EPHA3* | *EPHA5* | *EPHA7* |
| *EPHB1* | *ERBB2* | *ERBB3* | *ERBB4* | *ERG* | *ESCO2* | *ESR1* | *ETNK1* | *ETV6* | *EZH2* |
| *FANCA* | *FANCC* | *FANCD2* | *FAS* | *FAT1* | *FBXO11* | *FBXW7* | *FGF19* | *FGF3* | *FGF4* |
| *FGFR1* | *FGFR2* | *FGFR3* | *FGFR4* | *FLCN* | *FLT1* | *FLT3* | *FLT4* | *FOXL2* | *FOXO1* |
| *FOXP1* | *FURIN* | *FYN* | *GATA1* | *GATA2* | *GATA3* | *GNA11* | *GNA12* | *GNA13* | *GNAQ* |
| *GNAS* | *GNB1* | *GRIN2A* | *GSK3B* | *H1-2* | *H2BC5* | *H3C2* | *H3C8* | *HDAC1* | *HDAC4* |
| *HDAC7* | *HGF* | *HIF1A* | *HIST1H1B* | *HIST1H1D* | *HIST1H1E* | *HIST1H2AC* | *HIST1H2AG* | *HIST1H2AL* | *HIST1H2AM* |
| *HIST1H2BC* | *HIST1H2BG* | *HIST1H2BJ* | *HIST1H2BK* | *HIST1H2BO* | *HLA-A* | *HNF1A* | *HRAS* | *ID3* | *IDH1* |
| *IDH2* | *IGF1* | *IGF1R* | *IGF2* | *IKBKE* | *IKZF1* | *IKZF3* | *IL7R* | *INPP4B* | *IRF1* |
| *IRF4* | *IRF8* | *IRS2* | *JAK1* | *JAK2* | *JAK3* | *JARID2* | *JUN* | *KDM5A* | *KDM5C* |
| *KDM6A* | *KDR* | *KEAP1* | *KIT* | *KMT2A* | *KMT2B* | *KMT2C* | *KMT2D* | *KMT5A* | *KRAS* |
| *KSR2* | *LCK* | *LMO1* | *LTB* | *MALT1* | *MAP2K1* | *MAP2K2* | *MAP2K4* | *MAP3K1* | *MAP3K13* |
| *MAP3K14* | *MAPK1* | *MAPK3* | *MCL1* | *MDM2* | *MDM4* | *MED12* | *MEF2B* | *MEN1* | *MET* |
| *MGA* | *MGAM* | *MITF* | *MLH1* | *MOB3B* | *MPEG1* | *MPL* | *MRE11* | *MSH2* | *MSH6* |
| *MTOR* | *MUTYH* | *MYC* | *MYCL* | *MYCN* | *MYD88* | *NBN* | *NCOR1* | *NCOR2* | *NCSTN* |
| *NF1* | *NF2* | *NFE2* | *NFE2L2* | *NKX2-1* | *NOTCH1* | *NOTCH2* | *NOTCH3* | *NOTCH4* | *NPM1* |
| *NRAS* | *NSD1* | *NT5C2* | *NTRK1* | *NTRK2* | *NTRK3* | *P2RY8* | *PAK7* | *PALB2* | *PARP1* |
| *PAX5* | *PBRM1* | *PCBP1* | *PDCD1* | *PDGFRA* | *PDGFRB* | *PDPK1* | *PDS5B* | *PHF6* | *PIGA* |
| *PIK3C2G* | *PIK3C3* | *PIK3CA* | *PIK3CG* | *PIK3R1* | *PIK3R2* | *PIM1* | *PLCG1* | *PLCG2* | *PMS2* |
| *PNRC1* | *POT1* | *PPP2R1A* | *PRDM1* | *PRKAR1A* | *PTCH1* | *PTEN* | *PTPN1* | *PTPN11* | *PTPN2* |
| *RAD21* | *RAD50* | *RAD51* | *RAD51B* | *RAD51C* | *RAD51D* | *RAD52* | *RAD54L* | *RAF1* | *RARA* |
| *RB1* | *REL* | *RET* | *RHOA* | *RICTOR* | *RNF43* | *ROBO1* | *ROS1* | *RPTOR* | *RRAGC* |
| *RTEL1* | *RUNX1* | *RUNX1T1* | *SAMHD1* | *SDHA* | *SDHB* | *SDHC* | *SDHD* | *SETBP1* | *SETD1A* |
| *SETD1B* | *SETD2* | *SETD3* | *SETD4* | *SETD5* | *SETD6* | *SETD7* | *SETDB1* | *SETDB2* | *SF3B1* |
| *SGK1* | *SH2B3* | *SMAD2* | *SMAD4* | *SMARCA4* | *SMARCB1* | *SMARCD1* | *SMC1A* | *SMC3* | *SMG1* |
| *SMO* | *SOCS1* | *SOX2* | *SP140* | *SPEN* | *SPOP* | *SRC* | *SRSF2* | *STAG1* | *STAG2* |
| *STAT3* | *STAT5A* | *STAT5B* | *STAT6* | *STK11* | *SUFU* | *SUZ12* | *SYK* | *TBL1XR1* | *TBX3* |
| *TENT5C* | *TERT* | *TET1* | *TET2* | *TET3* | *TGFBR2* | *TNFAIP3* | *TNFRSF14* | *TOP1* | *TP53* |
| *TP53* | *TP63* | *TRAF2* | *TRAF3* | *TRAF5* | *TSC1* | *TSC2* | *TSHR* | *TYK2* | *U2AF1* |
| *U2AF2* | *UBR5* | *VAV1* | *VAV2* | *VHL* | *WHSC1* | *WT1* | *XBP1* | *XPO1* | *ZRSR2* |
